# Lipid Fingerprinting by MALDI Biotyper Sirius Instrument Fails to Differentiate the Three Subspecies of the *Mycobacterium abscessus* Complex

**DOI:** 10.1101/2024.09.23.24314022

**Authors:** Mitsunori Yoshida, Hanako Fukano, Koji Yahara, Satoshi Nakano, Takeshi Komine, Masato Suzuki, Azumi Fujinaga, Kohei Dohke, Yoshihiko Hoshino

## Abstract

The number of patients suffering from *Mycobacterium abscessus* complex (MABC) pulmonary diseases is steadily increasing. MABC consists of three subspecies, and it is recommended that the three subspecies be distinguished because of their differing macrolide susceptibilities. Unfortunately, current methods are inefficient due to their high cost, complexity, and time requirements. The third-generation Bruker MALDI Biotyper (MBT) Sirius has the capability to detect phospholipids and glycolipids using negative-ion mode. Mycobacterial cell walls are rich in lipids, and if lipid structure diversity can serve as species-specific fingerprints, this method may provide an alternative for microbial identification. This study aimed to examine the accuracy of discriminating between the three MABC subspecies by lipid profiling. Our best model failed to differentiate the three subspecies. Even in the two-dimensional space of the most significant peaks, *M. abscessus* and *M. massiliense* could not be separated. The agreement rate between lipid fingerprint-based and WGS-based identification was, at most, 47.2% for negative-ion mode. Even after applying recent machine learning algorithms to detect variables and create predictive models, accuracy remained at 50%. Our results suggest that using lipid fingerprinting alone to differentiate the three MABC subspecies is currently inadequate. Further advancements and standardization of MALDI-TOF MS-based methods are necessary for the routine differentiation of MABC subspecies in clinical settings.

## Manuscript

The number of patients suffering from *Mycobacterium abscessus* complex (MABC) pulmonary diseases is steadily increasing. MABC consists of three subspecies and is often termed an “antibiotic nightmare” due to its resistance to most antibiotics. The 2020 ATS Clinical Practice Guideline recommends distinguishing MABC subspecies because of their differing susceptibilities to macrolides (1). Unfortunately, current methods, such as HPLC, multilocus sequence typing (MLST), or whole-genome sequencing (WGS), are inefficient due to their high cost, complexity, and time requirements. Matrix-assisted laser desorption ionization time-of-flight (MALDI-TOF) mass spectrometry (MS) has been widely used in laboratories as a cost-effective and accurate method for identifying mycobacterial species. However, distinguishing between MABC subspecies using proteomic profiling remains challenging (2–4).

The third-generation Bruker MALDI Biotyper (MBT) Sirius has enhanced capabilities, such as detecting phospholipids and glycolipids using negative-ion mode, in addition to the positive-ion mode of previous versions. Mycobacterial cell walls are rich in lipids, and if lipid structure diversity can serve as species-specific fingerprints, this method may provide an alternative for microbial identification. A recent study suggested that lipid profiling could differentiate the three MABC subspecies (5), but its accuracy has not been fully evaluated. This study aimed to examine the accuracy of discriminating between the three MABC subspecies by lipid profiling using a set of MABC isolates.

We used the same 148 clinical isolates described in our previous publication (6), for which we had performed WGS. Mass spectra were acquired using the MBT Sirius in negative-ion mode, and data analysis was conducted using ClinProTools 3.0 (Bruker). A discrimination model was developed based on 20 clinical isolates and 3 reference strains, covering the MABC phylogenetic tree (Figure S1). This model was then applied to 127 clinical isolates (Table S1). Additionally, machine learning models—random forest (RF), Comb_Fisher, Max_Vote, multinomial log-linear models, neural networks, support vector machine (SVM), and extreme gradient boosting tree—were employed using the MSclassifR package (v0.3.2, https://github.com/agodmer/MSclassifR_examples).

Our best model failed to differentiate the three subspecies, as they overlapped on the principal component planes (Figure A). Even in the two-dimensional space of the most statistically significant peaks (peaks 72 and 31), *M. abscessus* and *M. massiliense* could not be separated (Figure B). The agreement rate between lipid fingerprint-based and WGS-based identification was, at most, 47.2% for negative-ion mode (Figure C). Even after applying recent machine learning algorithms to detect variables and create predictive models, accuracy remained at 50% (Table S2). Khor et al. employed ethanol for preparation and positive-ion mode for MS analysis, concluding that the wave pattern between 1200 and 1450 m/z could differentiate the three subspecies. However, we could not replicate their findings (5).

**Figure.**
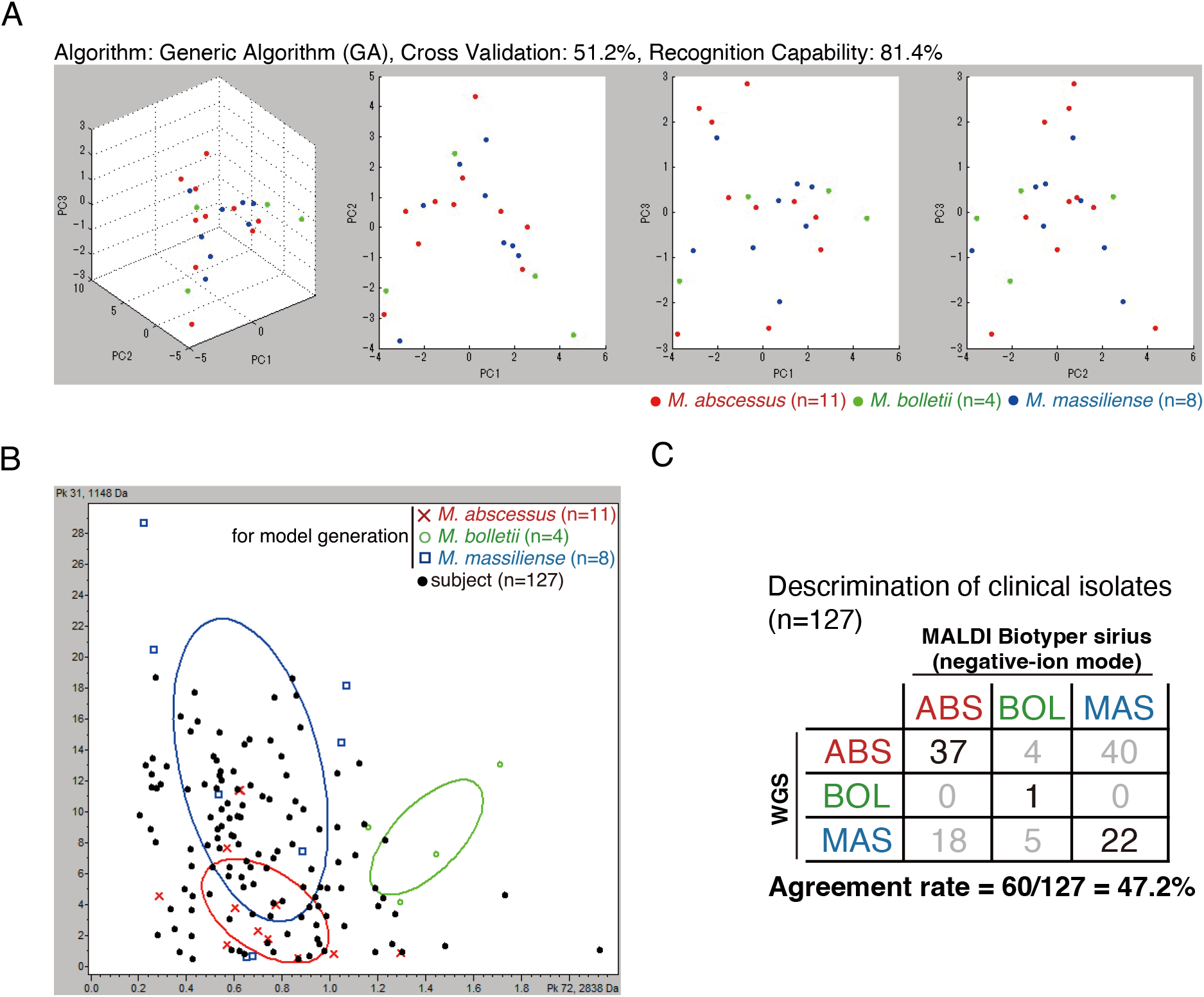
(A) Principal component analysis (PCA) of the peaks (760-3200 m/z) of *M. abscessus* complex. A scatter plot of the first three principal components (PC1 to PC3) of the peak lists of MBT sirius with negative-ion mode shows the distribution of the isolates of *M. abscessus* subsp *abscessus* (red, n=11), *M. abscessus* subsp. *massiliense* (blue, n=8), and *M. abscessus* subsp. *bolletii* (green, n=4). These 23 isolates, including three reference strains (ATCC 19977, JCM 15300, and BD), cover the genomic diversity of the MABC (arrows in Fig. S1). (B) A scatter plot of the two peaks of MBT sirius with negative-ion mode that are statistically most significant peaks to identify MABC subspecies (peaks 72 and 31) are shown. A set of MABC clinical isolates (black, n = 127) were coplotted onto the two-dimensional space between these peaks. (C) Agreement rate between subspecies identification using WGS data and MBT sirius data. Classification of clinical isolates (n = 127) was performed using a model generated with ClinProTools 3.0 (GA, same model as A). Subspecies identification using WGS data was performed as described previously (6).

Our results suggest that using lipid fingerprinting alone to differentiate the three MABC subspecies is currently inadequate. A recent study successfully differentiated the subspecies using machine learning analysis of proteomic profiling (acquired in positive-ion mode with the MBT Sirius) (7). However, this method has not yet been integrated into the commercial software offered by Bruker, and its implementation in clinical settings will take time. Furthermore, in our study, lipid profiling data did not show improved accuracy using machine learning methods. Therefore, to rapidly and simply distinguish the three MABC subspecies in clinical practice, alternative methods, such as the recently developed DNA chromatography kit (6) or the Genotype NTM-DR (8), are required. Further advancements and standardization of MALDI-TOF MS-based methods are necessary for the routine differentiation of MABC subspecies in clinical settings.

## Data Availability

All data produced in the present study are available upon reasonable request to the authors. All raw read data for sequenced strains in this study were deposited in the DNA Data Bank of Japan (DDBJ) and mirrored at the National Center for Biotechnology Information (NCBI) under BioProject accession number PRJDB10333.

## Ethics Statement

As described elsewhere (6), the National Institute of Infectious Diseases medical research ethics committee reviewed and approved this study, which included human subjects (#1004 and #1005 for the Japanese and Taiwanese, respectively).

## Acknowledgment

The authors thank Akiko Yamashita, Yukari Nogi, and Ginko Kaneda, Department of Mycobacteriology, National Institute of Infectious Diseases, for their assistance. This study was in part supported in part by grants from the Japan Agency for Medical Research and Development (AMED) to MY (JP24wm0225022 and JP24fk0108714) and to YH (JP23fk0108608, JP24fk0108673, JP24gm1610003, JP24gm1610007, JP24wm0125007, JP24wm0225022, JP24wm0325054, and JP24fk0108701). This study was also supported in part by grants from the Japan Society for the Promotion of Science (JSPS) for International Collaborative Research to YH and MY (JP63KK0138), and for Scientific Research (A) to YH (JP24H00331), and for Scientific Research (C) to MY and YH (JP23K07665 and JP23K07958).

## Figure legends

**Supplementally figure.**
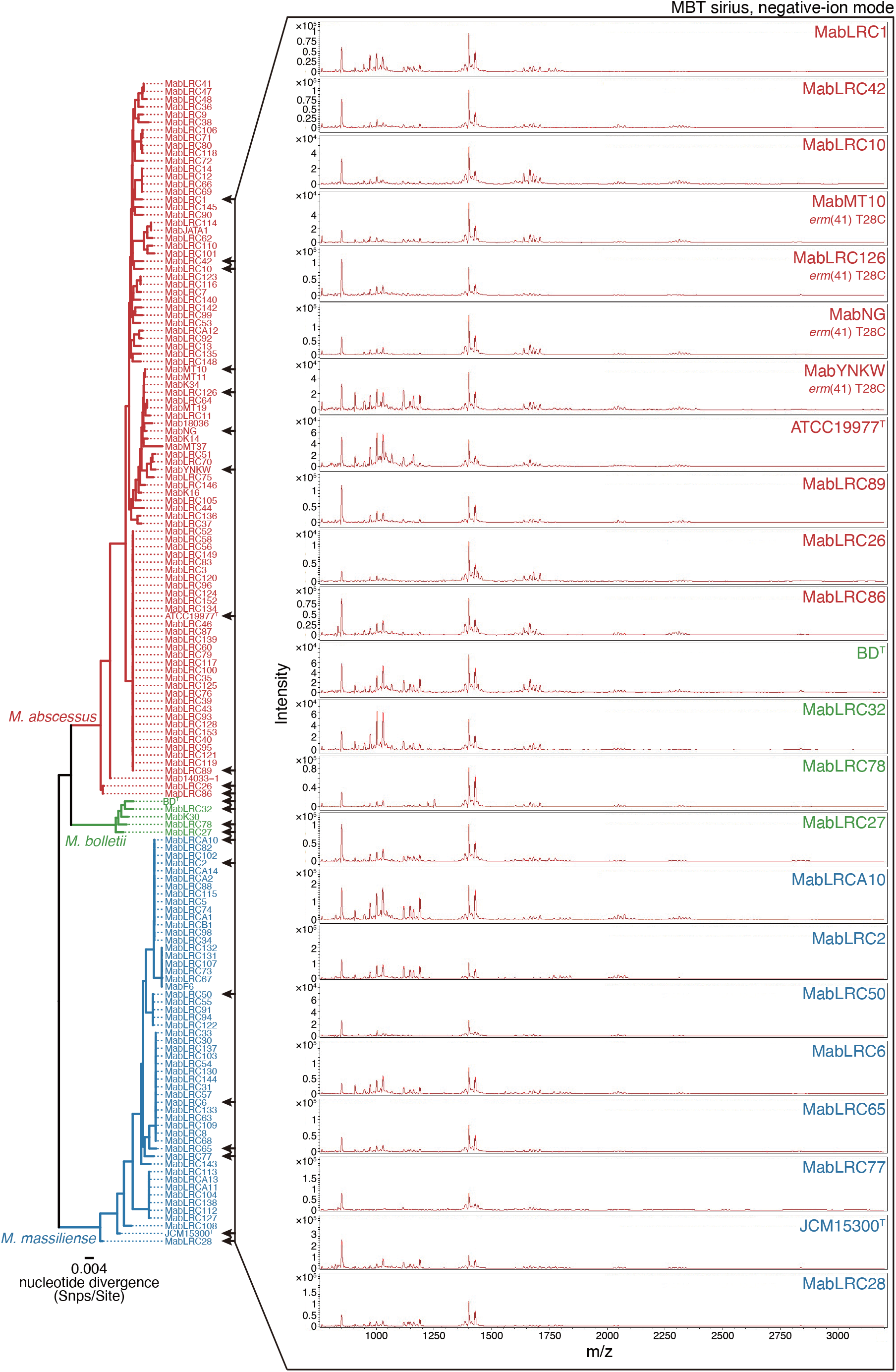
Maximum likelihood core-gene phylogeny of MABC isolates. Core gene alignment of 148 isolates and three reference strains (*M. abscessus* subsp *abscessus* ATCC19977, *M. abscessus* subsp. *massiliense* JCM 15300, and *M. abscessus* subsp. *bolletii* BD) of MABC was generated as described previously (6). The scale bar indicates the mean number of nucleotide substitutions per site (Snps/Site) on the respective branch. Samples are highlighted based on inclusion in three major clusters corresponding to MABC subspecies. Arrowheads indicate MABC isolates used for generating classification models. Mass spectra were acquired using MBT sirius with negative ion mode of MABC isolates and reference strains.

